# Effectiveness of a positive deviant intervention to improve appropriate feeding practices and nutritional outcomes in West Omo Zone, Maji District: South West Region, Ethiopia: A study protocol for a cluster randomized control trial

**DOI:** 10.1101/2022.03.17.22272474

**Authors:** Abraham Tamirat Gizaw, Pradeep Sopory, Morankar Sudhakar

## Abstract

**Background:** Non-optimal infant and young child feeding practices (IYCFP) are linked to malnutrition and infant mortality in poor countries, notably in Ethiopia. The majority of growth stalls occur within the first two years of life; hence, there is a need to discover interventions that enhance appropriate IYCFP for improving nutritional outcomes during this critical period. Using the experience of mothers who have come up with solutions to their IYCFP problems to educate others is is a potential pathway to initiate and sustain behavioral changes in resource-limited areas. However, such interventions are not widely implemented in Ethiopia.

**Objective:** This study aims to assess the effectiveness of a positive deviant (hearth nutrition education) intervention to improve appropriate feeding practices and nutritional outcomes in West Omo Zone, Maji District: South West region, Ethiopia.

**Methods:** A cluster randomized controlled trial will be conducted to compare the effect of positive deviant intervention versus routine health educationa. The intervention will be provided by positive deviant mothers who are members of the community using WHO infant and young child feeding guidelines, and “training of the trainers manual on counseling and supporting non-positive deviant mothers, infant and young child feeding” in the local language. Culturally appropriate and scientifically acceptable operational packages of information will be developed. Using preset criteria, 516 mothers will be recruited from 36 zones. The intervention arm will receive 12 demonstration (hearth) session in groups and every 15^th^ day home visit by positive deviant mothers. Data will be entered into epi-data version 3.1 and analyzed using STATA version 13.0. All analyses will be done as intention-to-treat. We will fit mixed effects linear regression models for the continuous outcomes and mixed effects linear probability models for the binary outcomes with the study zone as random intercept to estimate study arm difference (intervention vs. routine health education) adjusted for baseline value of the outcome and additional relevant covariates.

The protocol was developed in collaboration with the West Omo Zone and Maji Woreda Health Office. Ethical approval (Ref no: IHRPG/938/2020) was obtained from Jimma University, Institute of Health Research and Postgraduate Office. This study is funded by Jimma University research and postgraduate office.

**Discussion:** We expect that the trial will generate findings informing IYCFP and nutritional policies and practices in Ethiopia.

**Trial registration:** Registry: PACTR202108880303760 (30/08/2021); www.pactr.org, URL : https://pactr.samrc.ac.za/TrialDisplay.aspx?TrialID=16081

## Introduction

The Global Strategy for Infant and Young Child Feeding Practices (IYCFP) provides a framework for enhancing health, noting the need to review policy and program implementation to identify gaps and take action to bridge them [1]. IYCFP has the largest potential influence on child survival of any established preventive health and nutrition intervention framework. Malnutrition in children mostly occurs during the first two years of life due to a high demand for nutrients to sustain rapid growth and development; hence, the first two years of life are seen as a key window of opportunity to reduce malnutrition. The World Health Organization (WHO) suggests three IYCF indicators for children aged 6–24 months: minimal dietary diversity (MDD), minimum meal frequency (MMF), and minimum adequate diet (MAD) [2, 3]. The period between 6 and 24 months of age is a time of nutritional vulnerability because nutrients, particularly micronutrients, and energy supplied only from breast milk will not be adequate to fulfill the child’s needs. A key worldwide health issue is ensuring appropriate nutrition between the ages of 6 and 24 months [4-6].

In 2010, around 104 million children under the age of five were underweight and 171 million were stunted worldwide. Around 90 percent of stunted children reside in 36 countries, and children under the age of two are the most vulnerable to malnutrition. Malnutrition accounts for around 5.6 million of the 10 million child deaths each year, with severe malnutrition accounting for approximately 1.5 million of these fatalities [7,8]. Every day, between 3000 and 4000 infants die in poor countries as a result of diarrhea and severe respiratory infections caused by a lack of breast milk. In Ghana, only around 12.0% of young children are fed with infant feeding bottles [9,10]. While undernutrition (low weight for age) is widely recognized, the relevance of acute malnutrition is rarely, if ever, emphasized. This is a significant omission; acute malnutrition is a very prevalent disease with high rates of death and morbidity that need specific treatment and preventative measures [11]. Maternal low knowledge, negative attitude, low self-efficacy, and cultural influences and taboos all seem to have a significant impact on mothers’ infant-feeding practices and their children’s eating patterns [12,13]. Studies conducted in different parts of the world such as Brazil and Indonesia have revealed that knowledge, culture, self-efficacy, and beliefs are strong predictors of IYCFP [14,15]. Another study conducted in Turkana in Africa, also found that although mothers were aware of the length of exclusive breastfeeding, with 85.6 percent reporting that EBF should last 6 months, most were were unaware of the need for continuing breastfeeding beyond to the age of 24 month [16].

Sub-Saharan Africa (SSA) has one of the lowest breastfeeding rates, with 37 % of infants aged less than six months being exclusively breastfed. Lower proportions of infant feeding practices have been recorded in several SSA nations, where diarrhea is still a major cause of morbidity and mortality. Lower socioeconomic level, home childbirth, culture, and inadequate implementation and monitoring of programs are all plausible causes for unsatisfactory exclusive breast feeding practices in Africa [17-20]. The Ethiopian government developed and implemented the IYCFP guideline in 2004 to improve feeding practices and made different agreements to reduce infant mortality that results from malnutrition, including the Sekota agreement that aimed for a 1,000-day Nutrition Service program [21]. However, children’s malnutrition remains a significant public health challenge in Ethiopia.

According to the available research, in Ethiopia, poor dietary practices among children reported being malnourished ranged from 15.2–35.5 %. According to research conducted in Shashemene, 215 (65.7 percent) of children aged 6 to 24 months began solid, semi-solid, and soft meals between the ages of 6 and 8 months. Only 128 (39.1 %) of infants aged 6 to 24 months fulfilled the criteria for minimum dietary diversity (MDD), which is the consumption of four or more food groups from the seven food groups [22]. The percentage of stunted and underweight children is greater in rural areas than in urban ones, reflecting poor childcare practices in rural areas. Eighty-five percent of children scored poorly on dietary diversity [23, 24]. According to a study conducted in the Jimma zone on complementary feeding practices, the majority (88.9 %) of the children were exclusively breastfed, and 75.6 % were breastfed up to the age of two years [3].

The goal of this study is to use the positive deviant (hearth nutrition education) intervention (PDI) to improve appropriate IYCFP and nutritional outcomes. The positive deviant approach (PDA) provides a successful framework for enhancing child feeding, caring, and health-seeking behaviors for all community members, even the extremely poor [25]. In Ethiopia, only a few behavioral change interventions have been conducted with the aim of improving infant and young child feeding practices [26,27]. The reports of these projects focus either on implementation fidelity or implementation research. Moreover, none of the interventions were provided by positive deviant mothers using both demonstrations (hearth session) and home visits for an extended period in order to improve infant and young child feeding practices.

The aim of the planned trial is to examine the effect of positive deviant intervention to improve breastfeeding knowledge, attitude, self-efficacy, complementary feeding behaviors, and anthropometric outcomes in a cluster randomized community based behavioral promotion trial. The intervention will focus on intensively sparking local awareness and behavioral change, making the community to be empowered by appropriate feeding practice to enhance the health of infants and young children.

## Methods

### Design

A cluster-randomized controlled single-blinded parallel groups, two arms, trial with 1:1 allocation ratio was designed to examine positive deviant intervention (PDI) provided for mothers’ to improve IYCFP, nutritional outcomes, knowledge, attitude, self-efficacy, and health-seeking behaviors of mothers’ toward their infants and young children. The description of the study protocol conforms to the specifications of the Standard Protocol Items Recommendations for Intervention Trials (SPIRIT) checklist displayed in fig 1.Cluster randomized control was chosen to avoid contamination among treatment groups. Clusters are zones, i.e. small administrative units found in Maji Woreda, West Omo Zone (see fig 2).

**Fig. 1.**
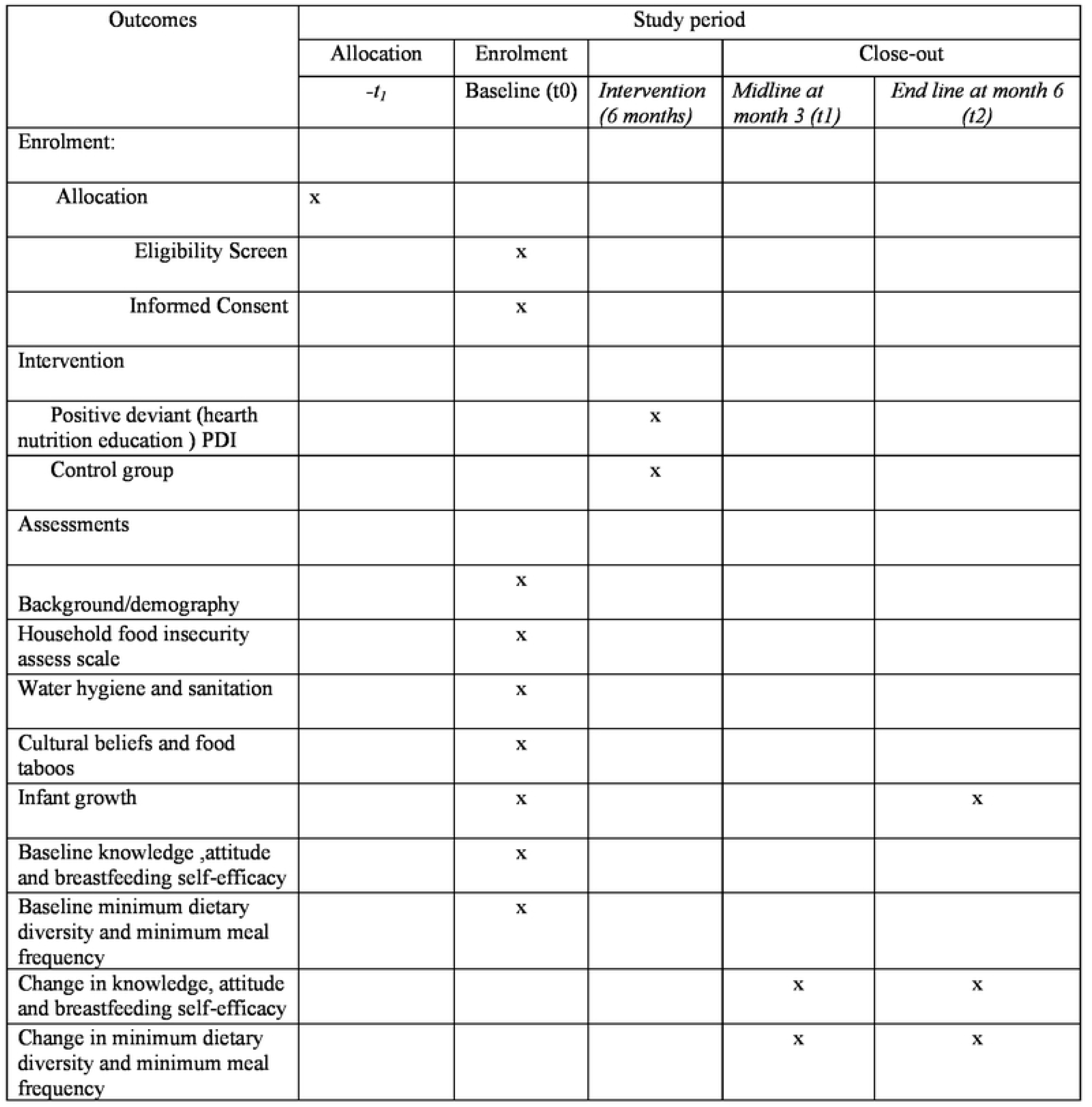
Standard Protocol Items: Recommendation for Intervention Trials (SPIRIT)

**Fig. 2.**
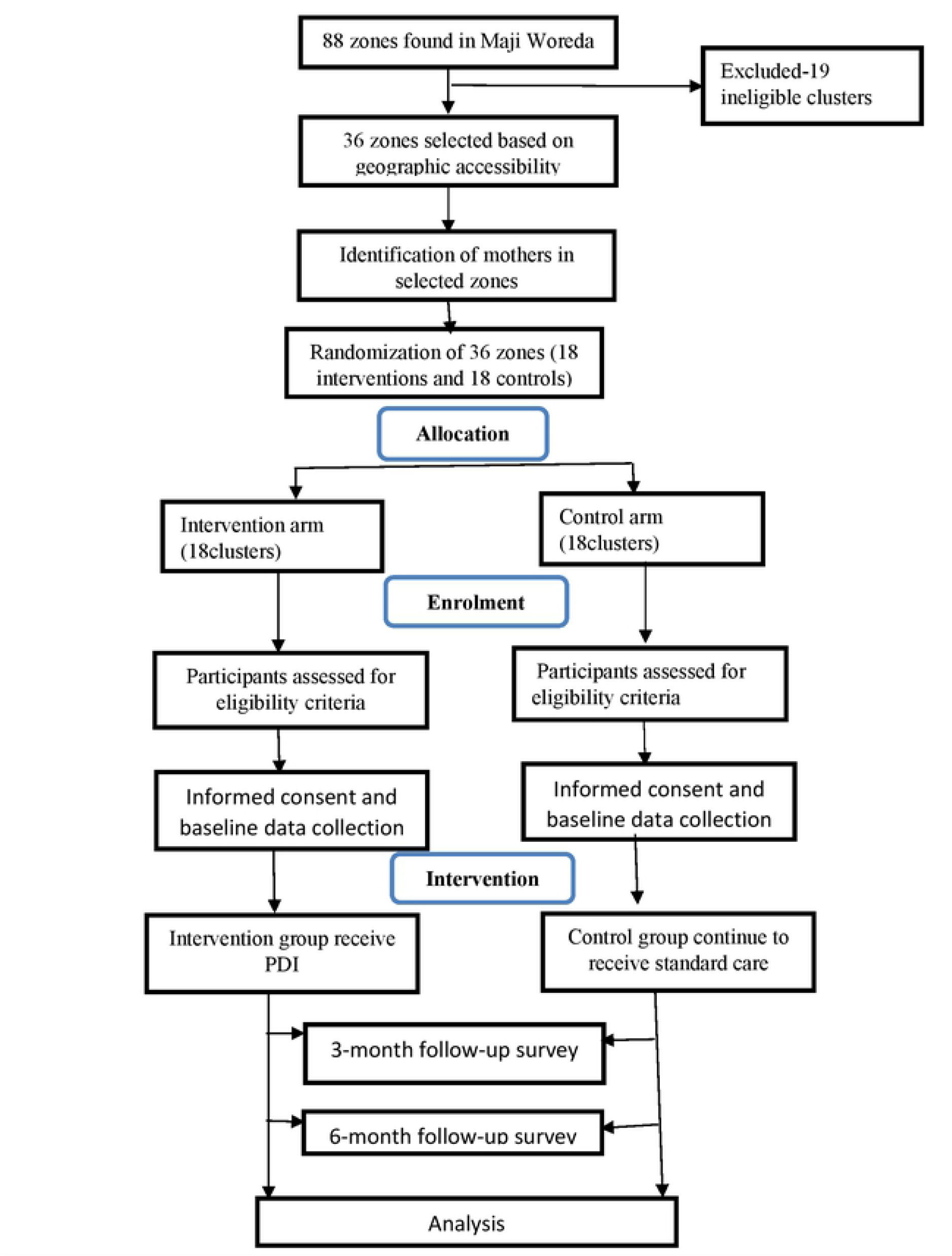
Flow of participants.

### Setting

This study will be conducted in the Maji district, West Omo zone, in Southern Ethiopia. Maji is one of the districts in the South West region. Maji is bounded on the south by the Kibish River, which divides it from South Sudan, on the west by Surma, on the northwest by Bero, on the north by Meinit Shasha, and on the east by the Omo River, which separates it from the Debub Omo Zone. There are 117 kebeles in the zone, with a population of 1,272,943. The Maji district has a total of 22 kebeles. In the Maji district, there are a district hospital, four health centers, and 22 health posts. Maji district is semi-pastoralist, with the bulk of the people relying on traditional rain-fed agriculture and rearing cattle. Maji district has a population of 230,777 people and is located 817 kilometers from Addis Ababa, Ethiopia [28]. The trial will take place in the Maji district, one of the districts of the West Omo Zone. The district is split into kebeles, which are the lowest administrative units, and each kebele is divided into four small zones (see fig 3).

**Fig. 3.**
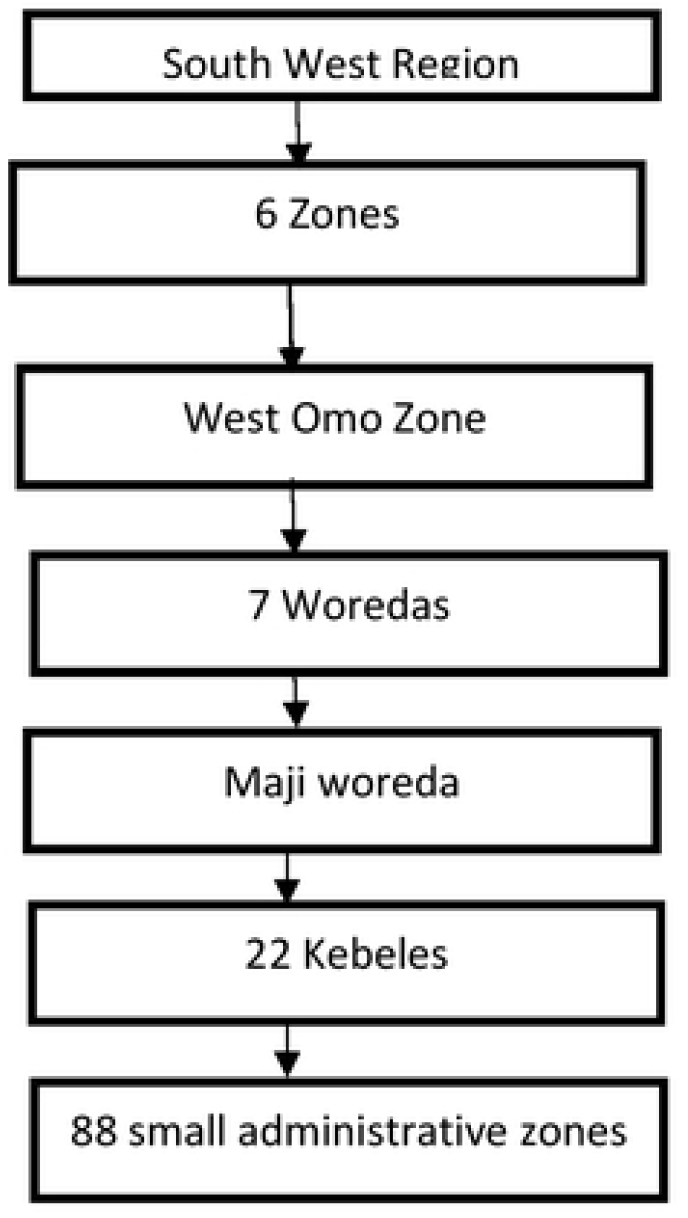
Flow diagram of structures in South West region, West Omo Zone, Maji Woreda

### Eligibility criteria for clusters

Out of 88 zones found in Maji Woreda, 36 clusters that are not adjacent to each other and are geographically accessible will be selected randomly for the study, resulting in 18 interventions and 18 controls (Fig 2).

### Eligibility criteria for the participants

The study will comprise Maji district mothers and their infants and young children (IYC) age 0-24 month. Inclusion criteria will be mothers living in the selected clusters with no plan to move away during the intervention period, without psychiatric illness, capable of giving informed consent, and willing to be visited by supervisors and data collectors. Infants and young children with no severe malnutrition and no severe illness will be included. Exclusion criteria will be mothers with a severe psychological illness which will interfere with consent and children with severe illness or clinical complications, which will potentially influence the study outcomes will excluded.However, such children will be refered to the nearby health institution for better care.

### Sample Size Determination

The sample size was calculated using the sample size formula for cluster randomized control trials. The following formula was used to calculate sample size with the following assumption to improve appropriate IYCFP from 7% to 14 % [29], at 95% confidence interval (CI), 80 % power, assuming an intra-correlation coefficient (ICC) of detectable differences in IYCFP indicators in Ethiopia which is 0.03 [30].

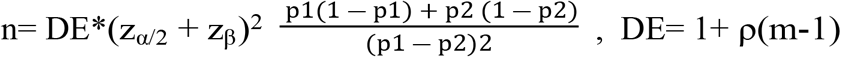

*Where:*

*p1: the proportion of outcome from the control group= 0*.*07, p2: the proportion of expected outcome from the intervention group =0*.*14,α :level of significant =0*.*05, Power (1-β)=0*.*8, Z-Alpha value (Z* _*α*/2_*)=1*.*96, Z-Beta value (z*_*β*_*)=0*.*84, ρ* : *intracluster correlation=0*.*03, m=number of mothers in each cluster =12, DE: design effect, calculated using the following formula =* DE= 1+ *ρ(m-1)=1*.*33 and n=required sample size. Assuming the number of mothers in each cluster is 12*.

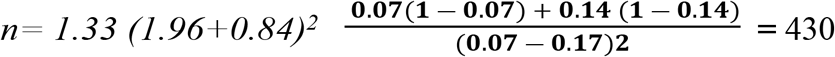

Adding 20% of the sample size for lost to follow-up, the final sample size is 516 mothers (258 from the intervention arm and 258 from the control arm). For a cluster size of 12, it was calculated that we need 36 clusters/zones.

### Sampling and randomization procedures

Zones in the kebeles will form the unit of randomization for the trials, while mothers within the zones form the unit of observation. From 7 woredas in the West Omo zone, Maji Woreda was selected purposively. After identifying and listing the 88 zones found in Maji Woreda, 36 non-adjacent zones will be selected. Then eligible mothers will be identified from the selected zones using the health extension worker’s logbook before the zones are randomized either into treatment or control groups. Simple randomization with 1:1 allocation will be used to randomize the zones to either the control or the interventions arms. First, the 36 zones will be listed alphabetically and then a list of random numbers will be generated in MS Excel 2016 and the generated list will be fixed by copying them as “value “next to the alphabetic list of zones. These will be arranged in ascending order according to the generated random numbers. Finally, the first 18 zones will be selected as intervention clusters and the last 18 as control clusters (Fig 2).

### Intervention

#### Positive deviance (hearth nutrition education) perspective

The positive deviant (hearth nutrition education) approach identifies and transfers good behaviors done by mothers of well-nourished children from disadvantaged households to others in the community with malnourished children [31]. The approach identifies good child feeding, care, and health-seeking behaviors that are available to all community members, even the extremely poor [32]. Positive deviant mothers will be selected based on a community-wide meeting with village leaders, women representatives in the community, and women health development army leaders working together with health extension workers. Mothers with young children who are found to be well-nourished will be identified. Additionally, the following criteria will be applied for the selection of positive deviant mothers (hearth nutrition educators): poor families, normal nutritional status, family who is representative of geographical and social groups living in the village, no severe health problems, belong to the community, and head of the household should have the same occupation as the majority of villagers. The positive deviant child cannot be: a big baby who is losing weight now, be a first-born or only child since it may receive special care, have any severely malnourished sibling, have any serious or typical social or health problem, have families enrolled in a supplementary feeding program and be a very small, low-weight baby who is now growing well [33]. Therefore, all the above-listed criteria for both mother and child should be well understood before deciding they are as positive deviant (hearth educator) mothers or not. Once the positive deviant mothers are selected then the selected positive deviant mothers (hearth nutrition educators) will be trained on UNICEF adopted guidelines for IYCFP [34].

The hearth nutrition education sessions will be provided for the selected non-positive deviant mothers in the intervention for 12 continuous days in groups to their nearby setting in the community. The hearth nutrition education elements such as knowledge, attitude, self-efficacy, appropriate feeding practice, hygiene, and health-seeking behavior. The positive deviant mothers will be assigned to the non-positive deviant (NPD) mothers and make regular visits to the household and provide information on exclusive breastfeeding, complementary feeding (MMF and MDD) practice, caring, hygiene, health-seeking at the time of child illness, and then wards, nutrition session, on how to cook and feed locally available food items to their children through follow-up visits for every two weeks to the intervention arms (overall 12 times during the intervention period) [35]. The control arm will receive routine nutrition education from community health workers (Health Extension workers). A range of activities will be done simultaneously. Developing IEC materials and messages promoting IYCFP such as leaflets, and posters, that have been produced based on the local context. Lastly, the outcome data will be conducted to evaluate and compare the intervention and control arms changes in appropriate IYCFP and nutritional outcomes. The overall hearth nutrition educational lesson will target undernutrition to promote positive IYCFP. This study expects children of mothers who received nutrition education through the positive deviance approach to be significantly better nourished than their counterparts in the control condition.

### The intervention description

#### Control arm

mothers in the control group will receive the routine health and nutrition education provided by health extension workers (HEWs) working in their kebeles and zones.

#### Intervention arm

mothers in the intervention arm will receive positive deviant (hearth nutrition education) intervention by selected positive deviant mothers. The intervention is composed of the following elements: a) breastfeeding education to raise knowledge, attitude, and breastfeeding self-efficacy, b) complementary feeding support, c) counseling on how to increase consistency, quantity, and frequency of foods, using locally available foods, and d) practical demonstration how to cook locally available food items, counseling, and support by positive deviant mothers.

After being trained, positive deviant mothers will provide infant and young feeding education and support to the selected non-positive deviant mothers. Besides the routine information and education provided to the mothers, each visit will be designated to cover a specific topic related to the outcome of the study.

### Nutrition education for selected mothers

The intervention arm will receive IYCFP and nutrition education with standard manuals prepared using the local language. During each visit, positive deviant mothers will cover the details of the importance of breastfeeding, complementary feeding, and feeding an ill child. The discussion will combine the use of information education and communication (IEC) material and practical demonstration on proper child feeding (breastfeeding and complementary feeding). Mothers will be encouraged to ask any questions related to the topic discussed.

Positive deviant mothers will use culturally appropriate language in the form of a poster to illustrate the new information (eg. correct and incorrect breastfeeding, preparation of the enriched flour, appropriate consistency (thickness) and inappropriate consistency of complimentary food, the importance of significant others support) and the benefit of applying the recommended infant and young child feeding practices (pictures of the babies who were appropriately fed versus those who were not).

#### Every fifteen days visits

positive deviant mothers will visit non-positive deviant mothers every 15^th^ days, with each mother visited twice a month for education and demonstration. During each visit, mothers will be observed breastfeeding, appropriate feeding, and preparation for complementary feeding provided, solving any breastfeeding problems, inappropriate feeding, dietary diversity, required consistency, and hands-on guidance when necessary. They will support and encourage the mothers to follow the appropriate infant and young child feeding practice from 0-24 months. Positive deviant mothers will also promote personal and domestic hygiene, such as handwashing before feeding, after toilet, and changing babies’ diapers.

#### Supervisors

Three persons will be involved in the supervision of the positive deviant mothers. The supervisors’ main responsibility will be to provide supportive supervision and monitor the positive deviant mothers. Supervisory visits will be conducted by the researcher along with the supervisor every fifteen days. Positive deviant mothers will receive feedback on their work from the supervisors during monthly supervision meetings.

### Study Objectives/hypotheses

#### Research hypotheses

Positive deviant (hearth nutrition education) intervention arm will significantly improve appropriate feeding practices and nutritional outcomes among infant and young children 0-24 months compared with the control arm.

#### Primary Objective

This study has two primary objectives:

First, at baseline, midline, and endline, this study will compare the effectiveness of a positive deviant (hearth nutrition education) intervention versus routine nutrition-related health education provided by health extension workers (community health workers) on mothers’ knowledge, attitude, and self-efficacy regarding IYCFP. Second, the nutritional outcomes in the control and intervention arms will be compared at baseline, and 6-month follow-up.

#### Secondary Objectives

This study also has one secondary objective, which is to explore barriers and facilitators to IYCFP at baseline through a qualitative study to supplement the quantitative study.This will help to understand the cultures and other factors influencing IYCFP in the study area.

### Outcome measurements

#### Primary outcomes

**Breastfeeding knowledge, attitude, and self-efficacy** is measured as breastfeeding knowledge score, attitude score, and breastfeeding self-efficacy score of mothers with index child from 0-24 months.

**Complementary feeding** assessed using the key indicators recommended by WHO [36], which include the timely introduction of solid, semi-solid, or soft foods, minimum dietary diversity, minimum meal frequency, and minimum acceptable diet calculated for the age range 6–11, 12– 17 and 18–24 months of age based on a

**Infant growth** WHO child growth standards (2009) will be used to estimate anthropometric status [36]: weight-for-length z-score (WLZ), length-for-age z-score (LAZ), and weight-for-age z-score (WAZ). Children who have WLZ below- 2 (WLZ < - 2) will be considered wasted, those with LAZ below- 2 (LAZ < - 2) stunted, and those with WAZ below - 2 (WAZ < - 2) underweight.

#### Secondary outcomes

Barriers and facilitatorsof IYCFP, health-seeking behavior, and morbidity status **(Table 2)**.

### Data collection tool and techniques

Ten data collectors and three suppervisors will be recruited and trained for three days. A strucued questionnaire prepared in Amharic for quantitative data collection and semi-structured questionnaire for qualitative data collection will be used to collect data. Components in the questionnaire will be prepared by adapting the validated tools for use in similar context. Data will be collected at baseline, 3^rd^ month, and 6^th^ month. Data on knowledge, attitude, self-efficacy, and complementary feeding will be collected at baseline, midline, and at study completion, whereas anthropometric measurements (length, weight and mid upper arm circumference (MUAC) will be done at baseline and at six month length (endline). Before each measurement, the weighing scale will be calibrated to zero. The weight and height/length measurements will be taken by standards [37]. Recumbent lengths for children aged 0-24 months will be measured to the closest 0.1 cm on a flat ground surface using a measuring tape and lying boards. Standing heights for older children will be measured to the closest 0.1 cm, with the head, shoulder, buttock, and heel all touching the stadiometer’s vertical surface. Measurements will be taken twice and the mean will be calculated.

All the data collectors will be trained on the content, questionnaire techniques, and measurements; in addition, produciblity and validity exercise will be conducted for the weight and length measurements. One of the requirement for the data collectors and suppervisors will be knowledge of local cultures and field study expertise. Breasfeeding knowledge, attitude and self-efficacy questionnaire will be developed and adapted using the following validated instruments: Breastfeeding knowledge [38], the Iowa Infant Feeding Attitude Scale (IIFAS) [39], and the short form of breastfeeding self-efficacy scale (BSES-SF) [40].

#### Breastfeeding knowledge

We used the breastfeeding knowledge questionnaire (BFKQ) consisting of 17 items to measure the knowledge of the participants about breastfeeding. There are three possible responses for each item (true, false and I do not know or not sure). Correct responses were scored as one, and zero for other options. Thus, the total scores ranged from 0– 17, these items were developed based on a study done among the Chinese mothers in English and translated to Amharic [41,42].

#### The Iowa Infant Feeding Attitude Scale (IIFAS)

consists of 17 items with a five-point Likert scale, rating maternal attitude towards breastfeeding translated from English. The scale ranging from strongly disagree to strongly agree on each item to indicate attitude towards infant feeding. A sum of scores ranging from 17 to 85 with the higher score reflecting a positive attitude whereas the lower score showing negative attitude. Attitude toward breastfeeding was categorized as follows: (1) positive to breastfeeding (IIFAS score 70-85), (2) neutral (IIFAS score 49-69), and (3) positive to formula feeding (IIFAS score 17-48) [43]. The IIFAS is a validated and reliable measure (Cronbach’s alpha scores ranges from (0.81–0.86) that evaluates breastfeeding attitudes in different cross-cultural settings [39,43,44]. Approximately half of the questions were negatively worded (i.e. 1, 2, 4, 6, 8, 10, 11, 14, and 17) [41,45].

#### The short form of breastfeeding self-efficacy scale (BSES-SF)

has been used widely with a variety of populations journals [46,47]. The overall score of the scale was calculated as the mean score of all items. A higher total score is indicative of a greater level of maternal breastfeeding self-efficacy. We used BSES-SF consisting of 14-items with a five-point Likert scale, developed to measure breastfeeding confidence in Amharic translated from validated English questionnaire from different studies, which measures the mother’s self-efficacy in her ability to breastfeed. All the items are preceded by the phrase “I can always “ and anchored with a 5-point Likert scale where 1 indicates not at all confident and 5-indicate always confident. All items are presented positively and scores are summed to produce a range from 14 to 70 [48].

### Data management

All filled questionnaire will be cheked for completeness by suppervisors and questionnaires with missing items will be returned to the data collectors for corrections. Mothers who are lost to follow up will be recorded along with their reasons.The following standard process will be implemented to improve the accuracy of the data enry and coding: double data entry and coding ;verification that the data is in the proper format and within an expected range of values; and independent source document verification of random subset of data to identify missing or apparently erroneous values. In order to ensure confidentiality, information about each zone and personal data of the partcipants will not be shared to any third party, both during and after the trial.

### Data prossessing and analysis

EpiData version 3.1 will be used for double data entry, and Stata version 16 (StataCorp) will be used for consistency checks and statistical analysis. Descriptive statistics will be utilized to assess and summarize sociodemographic, socioeconomic, child health status, child morbidity, and child feeding. The data for continuous variables will be given as a mean and standard deviation, or median and range, whereas categorical variables will be reported as a frequency and percentage. The t-test and analysis of variance will be used to compare group means for primary and secondary outcome variables. The Chi-square test will be used to examine the categorical variables. Child nutrition outcomes will be computed and compared to the WHO 2006 growth standards [49].

The results of group comparisons will be reported as a risk ratio for binary outcomes, equivalent to 2-sided 95 percent confidence intervals and related p values. All p-values will be scaled to two decimal places, with values less than 0.01 reported as < 0.01. Adjusted analyses utilizing baseline variables will be done using multivariate logistic regression to assess the ongoing effect of important baseline features on outcomes. For time-dependent variables such as morbidity, the Kaplan-Meier survival analysis will be employed. The intention-to-treat analysis will be utilized, and the clustering effect (using study zone as random intercept to account for clustering of subjects by zones) will be addressed. All analyses will be done with a 95% confidence interval. The significance level will be assigned at a p-value < 0.05.

For the qualitative data, all in-depth interviews and focus groups will be audio-recorded, which will be translated into English before being transcribed verbatim and thematically analyzed in ATLAS-ti version 9.1 using Systematic Text Condensation, a descriptive and exploratory analytic method [50].

### Ethics and Dissemination Plan

The study received ethical approval (reference number. IHRPG/938/20) from Jimma University’s Institute of Health Research and Postgraduate Office, Institutional Review Board. Administrative permission was acquired from Maji Woreda Administrative offices, and formal letters to the research area were obtained from Maji Woreda Health Office. Before enrolling in the study, study participants will provide informed verbal and written permission. Participants will be free to withdraw from the research at any time if they so choose. Anonymity and confidentiality will be guaranteed and upheld. To increase transparency, the complete protocol will be published in an open-access journal. However, to maintain confidentiality, the participant-level dataset and statistical code will not be made public. Any changes to the protocol will be reported to appropriate parties such as the trial registry and the ethics committee. The findings will be shared with internal, national, and international audiences via publishing in peer-reviewed journals and open access publications, and presentations at national and international conferences. Workshops will be offered as compensation for communities that participate in the project. The study’s findings will be presented in the West Omo Zone and Maji Woreda health offices. Furthermore, results will be presented in national and international conferences and workshops. Authorship eligibility guidelines and any intended use of professional writers. ICMJE guideline for authorship will be followed.

## Discussion

In the experimental arm, the PDI behavioral change communication approach will be provided concurrently to mothers to enhance their IYCFP and nutritional outcomes. To ensure accessibility and sustainability, the intervention is culturally adapted for implementation in the community context by selected PDM who solved their problems with locally accessible food items and brought about major changes in child feeding. Essentially, complementary feeding, which is MDD and MMF for young children, will be based on locally available and consumed food items in the community. It is expected that mothers will perceive child undernutrition as an important health concern and will be motivated to practice recommended actions to bring about positive behavioral changes. This study will improve knowledge and attitude, and the demonstration of how to cook and feed a child will improve skill acquisition, which, in turn, will enhance their self-efficacy. We predict our PDI will improve appropriate IYCFP from 7% to 14%, nutritional outcomes, knowledge, attitude, self-efficacy (KASe), and other cultural and gender issues influencing feeding practices as compared with routine nutritional education by health extension workers (community health workers). Finally, the findings will be scaled up to other regions and nationally to all of Ethiopia.

### Trial status

From the seven woredas (district) found in West Omo Zone, Maji woreda is selected purposely. Trainers’ and participants’ manuals were prepared in English and translated into Amharic, the language spoken locally. Information, education communication materials such as posters, leaflets, and visual teaching materials were developed in Amharic. This trial will be carried out from January 1,2022 to July 2, 2022.

## Data Availability

No datasets were generated or analysed during the current study. All relevant data from this study will be made available upon study

## Abbreviations

CF: Complementary feeding
EBF: Exclusive breastfeeding
IYCFP: Infant and young child feeding practice
IYCF: Infant and young child feeding
IYC: infant and young child
MAD: minimum adequate diet
MDD: minimal dietary diversity
MMF: minimum meal frequency
PDA: positive deviant approach
PDM: Positive deviant mothers
PDHNE-I: positive deviant (hearth nutrition education) intervention
NPD: non-positive deviant
SSA: Sub-Saharan Africa
UNICEF: United Nations Children’s Fund

## Authors’ contributions

**Conceptualization:** ATG, PS and MS.

**Data curation:** ATG, PS and MS.

**Formal analysis** : ATG.

**Funding acquisition** :ATG and MS.

**Investigators:** ATG, PS and MS.

**Methodology** : ATG, PS and MS.

**Project administration :** ATG, PS and MS.

**Suppervision:** ATG

**Visualization:** ATG,PS and MS.

**Writing-original draft:**ATG

**Writing-review and editing:**PS and MS

---

**Table 1.**
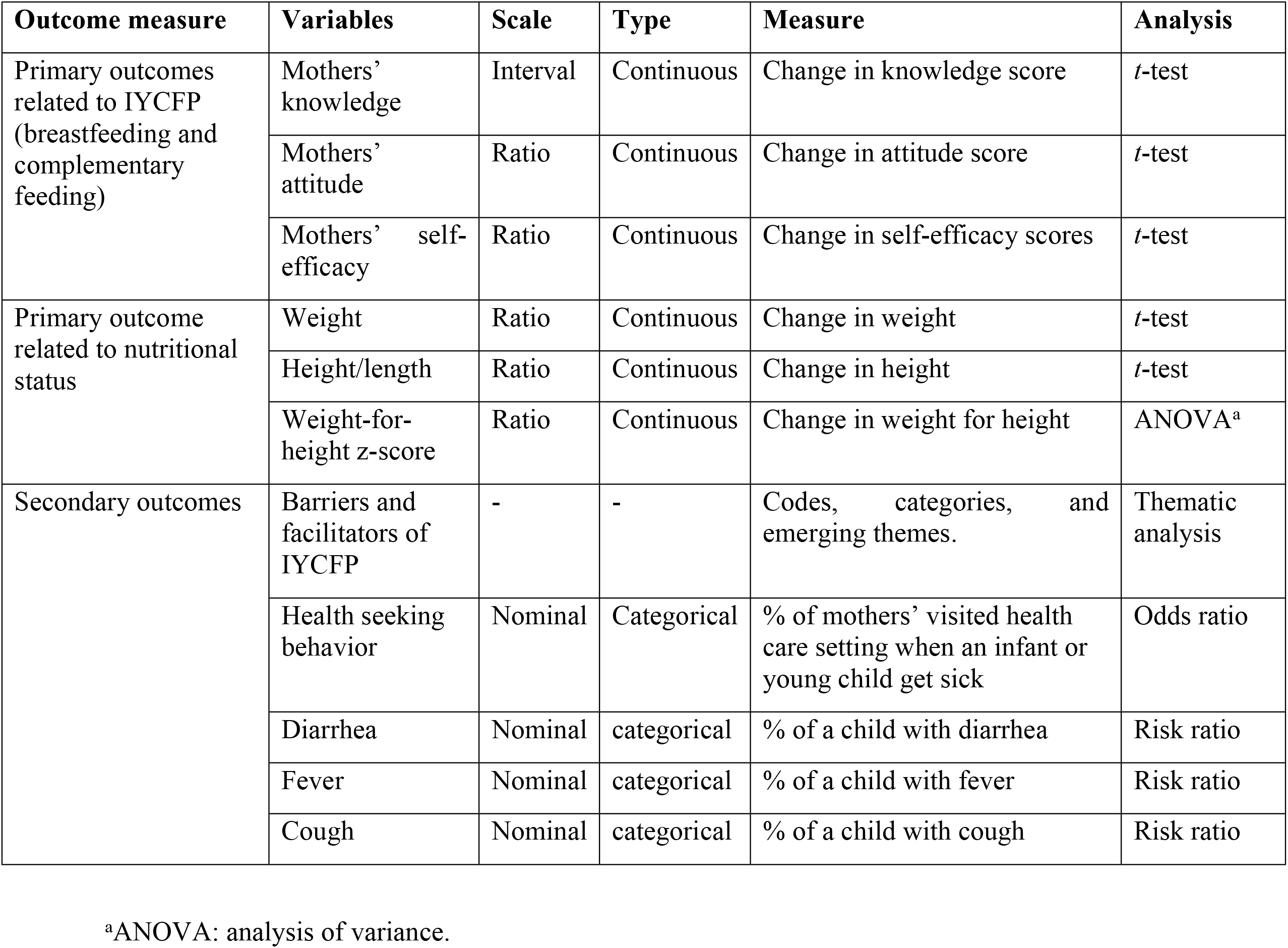
Primary and secondary outcome variables among study participants at baseline, 3 months, and 6 months

| Outcome measure | Variables | Scale | Type | Measure | Analysis |
| --- | --- | --- | --- | --- | --- |
| Primary outcomes related to IYCFP (breastfeeding and complementary feeding) | Mothers' knowledge | Interval | Continuous | Change in knowledge score | *t*-test |
|  | Mothers' attitude | Ratio | Continuous | Change in attitude score | *t*-test |
|  | Mothers' self-efficacy | Ratio | Continuous | Change in self-efficacy scores | *t*-test |
| Primary outcome related to nutritional status | Weight | Ratio | Continuous | Change in weight | *t*-test |
|  | Height/length | Ratio | Continuous | Change in height | *t*-test |
|  | Weight-for-height z-score | Ratio | Continuous | Change in weight for height | ANOVA<sup>a</sup> |
| Secondary outcomes | Barriers and facilitators of IYCFP | - | - | Codes, categories, and emerging themes. | Thematic analysis |
|  | Health seeking behavior | Nominal | Categorical | % of mothers' visited health care setting when an infant or young child get sick | Odds ratio |
|  | Diarrhea | Nominal | categorical | % of a child with diarrhea | Risk ratio |
|  | Fever | Nominal | categorical | % of a child with fever | Risk ratio |
|  | Cough | Nominal | categorical | % of a child with cough | Risk ratio |

<sup>a</sup>ANOVA: analysis of variance.

## References

1. Gupta A, Holla R, Dadhich JP, Suri S, Trejos M, Chanetsa J. The status of policy and programmes on infant and young child feeding in 40 countries. Health Policy Plan. 2013;28:279–298.

2. Reinbott A, Kuchenbecker J, Herrmann J, Jordan I, Muehlhoff E, Kevanna O, et al. A child feeding index is superior to WHO IYCF indicators in explaining length-for-age Z-scores of young children in rural Cambodia. Paediatr Int Child Health. 2015;35(2):124– 34.

3. Forsido SF, Kiyak N, Belachew T, Hensel O. Complementary feeding practices, dietary diversity, and nutrient composition of complementary foods of children 6 – 24 months old in Jimma Zone, Southwest Ethiopia. J Heal Popul Nutr. 2019;6(38):1–7.

4. Dagne AH, Anteneh KT, Badi MB, Adhanu HH, Ahunie MA, Aynalem GL. Appropriate complementary feeding practice and associated factors among mothers having children aged 6–24 months in Debre Tabor Hospital, North West Ethiopia, 2016. BMC research notes. 2019 Dec;12(1):1–6.

5. Quinn V, Zehner E, Schofield D, Guyon A, Huffman S. Maternal, Infant and Young Child Nutrition (MIYCN) Working Group: using the code of marketing of breast-milk substitutes to guide the marketing of complementary foods to protect optimal infant feeding practices. Maternal, Infant and Young Child Nutrition (MIYCN) Working Group: using the code of marketing of breast-milk substitutes to guide the marketing of complementary foods to protect optimal infant feeding practices. 2010.

6. Radwan H. Influences and Determinants of Breastfeeding and Weaning Practices of Emirati Mothers. 2011.

7. Ughade S, Chavan S, Narlwar U, Jadhao A, Adikane H. Cross sectional study of knowledge and practices regarding breast feeding amongst mothers belonging to tribal community in Melghat area, Amravati, Maharashtra, India. Int J Res Med Sci. 2017;5(3):990.

8. Collins S, Sadler K, Dent N, Khara T, Guerrero S, Myatt M. Key issues in the success of community-based management of severe malnutrition. 2013.

9. Babbel NF. Understanding child malnutrition in Ethiopia : Determinants of child caring practices, multiple anthropometric failures and seasonality of growth Understanding child malnutrition in Ethiopia : Determinants of child caring practices, multiple. 2017.

10. Chavan S, Jadhao A, Narlwar U, Ughade S, Adikane H. Cross sectional study of knowledge and practices regarding breast feeding amongst mothers belonging to tribal community in Melghat area, Amravati, Original Research Article Cross sectional study of knowledge and practices regarding breast feeding amongs. 2017;(July).

11. Lindsay AC, Machado MT, Sussner KM, Hardwick CK, Peterson KE. Infant-feeding practices and beliefs about complementary feeding among low-income Brazilian mothers: A qualitative study. Food Nutr Bull. 2008;29(1):15–24.

12. Arafat Y, Islam MM, Connell N, Mothabbir G, McGrath M, Berkley JA, et al. Perceptions of Acute Malnutrition and Its Management in Infants Under 6 Months of Age: A Qualitative Study in Rural Bangladesh. Clin Med Insights Pediatr. 2018;12:1–10.

13. Rakhee Verma PG. Impact Of Breast Feeding And Weaning Practices Associated With Morbidity In Rural Area Of Ghaziabad, Uttar Pradesh, India : A. Open Access J. 2015;6(4):4–7.

14. Lindsay AC, Wallington SF, Greaney ML, Hasselman MH, Tavares Machado MM, Mezzavilla RS. Brazilian Immigrant Mothers’ Beliefs and Practices Related to Infant Feeding: A Qualitative Study. J Hum Lact. 2017;33(3):595–605.

15. Awaliyah SN, Rachmawati IN, Rahmah H. Breastfeeding self-efficacy as a dominant factor affecting maternal breastfeeding satisfaction. BMC Nurs. 2019;18(Suppl 1):1–7.

16. Ochola S. Maternal Infant and Young Child Nutrition (Miycn) Knowledge, Attitudes, Beliefs and Practices (Kabp) Survey Report Turkana County. 2017.

17. Woolfenden S, Eastwood J, Page A, Agho K, Ogeleka P, Ogbo FA. Infant feeding practices and diarrhoea in sub-Saharan African countries with high diarrhoea mortality. PLoS One. 2017;12(2):e0171792.

18. Demilew YM. Factors associated with mothers’ knowledge on infant and young child feeding recommendation in slum areas of Bahir Dar City, Ethiopia: cross sectional study. BMC Res Notes. 2017;10(1):1–14.

19. Jones AD. Overcoming barriers to improving infant and young child feeding practices in the Bolivian Andes: the role of agriculture and rural livelihoods. 2011.

20. Gebre A, Surender Reddy P, Mulugeta A, Sedik Y, Kahssay M. Prevalence of Malnutrition and Associated Factors among Under-Five Children in Pastoral Communities of Afar Regional State, Northeast Ethiopia: A Community-Based Cross-Sectional Study. J Nutr Metab. 2019;2019:1–14.

21. Diddana TZ, Kelkay GN, Dola AN, Sadore AA. Effect of Nutrition Education Based on Health Belief Model on Nutritional Knowledge and Dietary Practice of Pregnant Women in Dessie Town, Northeast Ethiopia : A Cluster Randomized Control Trial. Hindawi J Nutr Metab. 2018;2018(1–10).

22. Yonas F, Asnakew M, Wondafrash M, Abdulahi M. Infant and Young Child Feeding Practice Status and Associated Factors among Mothers of under 24-Month-Old Children in Shashemene Woreda, Oromia Region,. Open Access Libr J Infant. 2015;39(2):1–15.

23. Kumera G, Tsedal E, Ayana M. Dietary diversity and associated factors among children of Orthodox Christian mothers / caregivers during the fasting season in Dejen District, North West. Nutr Metab (Lond). 2018;15(16):1–9.

24. Ashworth A, Ferguson E. Dietary counseling in the management of moderate malnourishment in children. Food Nutr Bull. 2009;30(3):405–33.

25. Doskey S, Mazzuchi T, Sarkani S. Positive Deviance Approach for Identifying Next-Generation System Positive Deviance Approach for Identifying Next-Generation System Engineering Best Practices. 2017;(December 2013).

26. Swanson V, Hart J, Byrne-Davis L, Merritt R, Maltinsky W. Enhancing behavior change skills in health extension workers in ethiopia: Evaluation of an intervention to improve maternal and infant nutrition. Nutrients. 2021;13(6):1–12.

27. USAID. Moving Nutrition Social and Behavior Change Forward. 2017. p. 1–12.

28. Teklehaymanot T, Giday M. Ethnobotanical study of wild edible plants of Kara and Kwego semi-pastoralist people in Lower Omo River Valley, Debub Omo Zone, SNNPR,. J Ethnobiol Ethnomedicine Res. 2010;6(23):2–9.

29. Ethiopia Demographic and Health Survey. 2016.

30. Moss C, Bekele TH, Salasibew MM, Sturgess J, Ayana G, Kuche D, et al. Sustainable Undernutrition Reduction in Ethiopia (SURE) evaluation study : a protocol to evaluate impact, process and context of a large-scale integrated health and agriculture programme to improve complementary feeding in Ethiopia. 2018;1–11.

31. Anino OC, Were GM, Khamasi JW. Impact evaluation of positive deviance hearth in Migori County, Kenya. Vol. 15, African Journal of Food, Agriculture, Nutrition and Development. 2016.

32. Nishat N, Batool I. Effect of “ Positive Hearth Deviance “ on feeding practices and underweight prevalence among children aged 6-24 months in Quetta district, Pakistan : A comparative cross sectional study. Sri Lanka J Child Heal. 2011;40(2011):57–62.

33. Sternin Monique, Sternin J, Marsh D. Designing a Community-Based Nutrition Program. Using the Hearth Model and the. Positive Deviance Approach - A Field Guide. Save Child. 1998;(December):1–89.

34. D’Alimonte MR. Qualitative Exploration of Behaviors Related to Positive Child Growth In an Urban Slum of Mumbai. ProQuest Diss Theses. 2014;(January):27.

35. Doskey S, Mazzuchi T, Sarkani S. Positive Deviance Approach for Identifying Next-Generation System Engineering Best Practices. Procedia Comput Sci. 2013;16:1112–21.

36. WHO. Growth velocity based on weight, length and head circumference Methods. 2009.

37. Cashin K, Oot L. GUIDE TO ANTHROPOMETRY A Practical Tool for Program Planners, Managers, and Implementers. 2018;

38. Hamze L, Mao J, Reifsnider E. Knowledge and attitudes towards breastfeeding practices: A cross-sectional survey of postnatal mothers in China. Midwifery [Internet]. 2019;74:68–75. Available from: https://doi.org/10.1016/j.midw.2019.03.009

39. Vijayalakshmi P, Susheela, D.M. Knowledge, Attitudes and Breast Feeding Practices of Postnatal Mothers : A Cross Sectional Survey. Int J Health Sci (Qassim). 2015;9(4):363– 72.

40. Monteiro JCDS, Guimarães CM de S, Melo LC de O, Bonelli MCP. Breastfeeding self-efficacy in adult women and its relationship with exclusive maternal breastfeeding. Rev Lat Am Enfermagem. 2020;28(2020):1–9.

41. Abdulahi M, Fretheim A, Argaw A, Magnus JH. Determinants of knowledge and attitude towards breastfeeding in rural pregnant women using validated instruments in ethiopia. Int J Environ Res Public Health. 2021;18(15).

42. Shohaimi NM, Mazelan M, Ramanathan K, Hazizi MSM, Leong YN, Cheong X Bin, et al. Intention and practice on breastfeeding among pregnant mothers in Malaysia and factors associated with practice of exclusive breastfeeding: A cohort study. PLoS One [Internet]. 2022;17(1 January):1–11. Available from: http://dx.doi.org/10.1371/journal.pone.0262401

43. Lau Y, Htun TP, Lim PI, Ho-Lim SST, Klainin-Yobas P. Psychometric Properties of the Iowa Infant Feeding Attitude Scale among a Multiethnic Population during Pregnancy. J Hum Lact. 2016;32(2):315–23.

44. Abdulahi M, Fretheim A, Argaw A, Magnus JH. Adaptation and validation of the Iowa infant feeding attitude scale and the breastfeeding knowledge questionnaire for use in an Ethiopian setting. Int Breastfeed J. 2020;15(1):1–11.

45. Inoue M. Breastfeeding and perceptions of breast shape changes in Australian and Japanese women. 2012.

46. Mohammadian M, Maleki A, Badfar G. Effect of continuous supportive telephone counselling on improving breastfeeding self-efficacy in mothers with late preterm infants four months after discharge: A randomized, controlled study. J Mother Child. 2021;25(1):44–50.

47. Guimarães CM de S, Conde RG, Gomes-Sponholz FA, Oriá MOB, Monteiro JC dos S. Factors related with breastfeeding self-efficacy immediate after birth in puerperal adolescents. Acta Paul Enferm [Internet]. 2017;30(1):109–15.

48. Tsaras K, Sorokina T, Papathanasiou I, Fradelos E, Papagiannis D, Koulierakis G. Breastfeeding Self-efficacy and Related Socio-demographic, Perinatal and Psychological Factors: a Cross-sectional Study Among Postpartum Greek Women. Mater Socio Medica. 2021;33(3):206.

49. WHO Child Growth Standards. Dev Med Child Neurol. 2009;51(12):1002–1002.

50. Malterud K. Systematic text condensation: A strategy for qualitative analysis. Scand J Public Health. 2012;40(8):795–805.

